# Natural immunity against COVID-19 significantly reduces the risk of reinfection: findings from a cohort of sero-survey participants

**DOI:** 10.1101/2021.07.19.21260302

**Authors:** Bijaya Kumar Mishra, Debdutta Bhattacharya, Jaya Singh Kshatri, Sanghamitra Pati

**Affiliations:** ICMR-Regional Medical Research Centre (Dept. of Health Research, Ministry of Health & Family Welfare, Govt. of India), Bhubaneswar-751023, Odisha, India

**Author notes:** **Address for correspondence : Dr. Sanghamitra Pati**, Scientist-G & Director, ICMR-Regional Medical Research Centre, (Dept. of Health Research, Ministry of Health & Family Welfare, Govt. of India), Chandrasekharpur, Bhubaneswar-751023.

**Keywords:** COVID-19, sero survey, natural immunity, sero-positive, sero-negative

## Abstract

Conflicting reports on the persistence of antibody levels in individuals recovered from COVID-19 infection, suggest that the immunity against COVID-19 may not be lasting for long. In India, by 30th June, 2021, not less than 30 million people were infected with COVID-19 and 0.39 million people were reported to have lost their life to the disease in India. I the current study we followed up with a subsample of our previous sero-survey participants to assess whether natural immunity against SARS-CoV-2 was associated with a reduced risk of re-infection. We conducted telephonic interview of a total of 3038 participants, out of which 2238 participants responded and 5 participants were found to be not alive, as conveyed by their close relatives. There was a non-response rate of 26.1%. Out of the 2238 participants, 1170 were sero-positive and 1068 were sero-negative for antibody against COVID-19. Our survey found that only 3 individuals in the sero-positive group got infected with COVID-19 whereas 127 individuals reported contracting the infection the sero-negative group. Interestingly, from the 127 sero-negative individuals who later contracted COVID-19 infection, 30 needed hospitalization, out of which 12 were on oxygen therapy, four in ICU and one was on ventilator. At the other hand, from the 3 sero-positives re-infected with COVID-19, one had hospitalization, but didnnot require oxygen support or critical care. These findings reinforce the strong plausibility that development of antibody following natural infection not only protects against re-infection by the virus to a great extent, but also safeguards against progression to severe COVID-19 disease.

## Introduction

We read with interest the communication by Poletti et al.,^1^ where the authors conclude that adherence to hygiene protocols and the maintenance of universal masking remain necessary ingredients in a multipronged strategy for infection reduction. Conflicting reports on the persistence of antibody levels in individuals recovered from COVID-19 infection, suggest that the immunity against COVID-19 may not be lasting for long.^2,3^ In India, by 30^th^ June, 2021, not less than 30 million people were infected with COVID-19 and 0.39 million people were reported to have lost their life to the disease in India.^4^ Three rounds of sero-survey in the state of Odisha have demonstrated an increasing trend of population level exposure to SARS-CoV-2.^5^ We followed up with a subsample of our previous sero-survey participants to assess whether natural immunity against SARS-CoV-2 was associated with a reduced risk of re-infection.

## Methodology

We conducted telephonic interview of a total of 3038 participants selected from our previous seroservey cohort from different district of Odisha. Data was recorded in preformed questionnaire. Consent was obtained prior to their enrollment in the study.

## Results & Discussion

Out of 3038 participants contacted over telephone, 2238 participants responded and 5 participants were found to be not alive, as conveyed by their close relatives. There was a non-response rate of 26.1%. Out of the 2238 participants, 1170 were sero-positive and 1068 were sero-negative for antibody against COVID-19. Our survey found that only 3 individuals in the sero-positive group got infected with COVID-19 whereas 127 individuals reported contracting the infection the sero-negative group. Taking into account a median follow-up duration of 258 days (66-319 days) and considering a CI of 95%, the incidence of COVID-19 among the sero-positive was 4.09 /1000 person-years [95% CI:0.84-11.94] while that among the sero-negative was 173.68 /1000 person-years [95% CI: 144.78-206.64], thus making a rate difference of -169.59 /1000 person-years (95% CI: -200.15 to - 139.03) and a rate ratio of 0.023 (95% CI: 0.007-0.073). This suggests that antibody produced in response to natural infection with COVID-19 is likely to reduce the risk of COVID re-infection by 97.7%. This is congruent with another similar study reporting that previous infection with COVID-19 decreases the risk of re-infection by 84%, with the median duration of protection being 7 months.^5^ Interestingly, from the 127 sero-negative individuals who later contracted COVID-19 infection, 30 needed hospitalization, out of which 12 were on oxygen therapy, four in ICU and one was on ventilator. At the other hand, from the 3 sero-positives re-infected with COVID-19, one had hospitalization, but didn’t require oxygen support or critical care.

## Conclusion

These findings reinforce the strong plausibility that development of antibody following natural infection not only protects against re-infection by the virus to a great extent,^6^ but also safeguards against progression to severe COVID-19 disease. However, further follow up must continue to draw concrete inferences on the protection conferred by naturally developing immunity after COVID-19 infection and its ability to protect against other emerging variants of the virus. Till that time, it is prudent to ensure adherence to COVID-19 preventive protocols in a large dense demography like India.

## Supporting information

Supplementary file for methodology

## Data Availability

Available on request to the corresponding author

## Data availability

All data in the manuscript are available on request to the corresponding author.

## Authors Contributions

SP designed the study. BKM, JSK & DB were responsible for data collection and analysis. SP, BKM, JSK & DB wrote the manuscript. All authors have read and approved the final manuscript.

## Acknowledgement

The authors gratefully acknowledge all the healthcare workers for their tireless dedication at each level to fight COVID-19 and for voluntarily participating in this cohort study. The authors are thankful to the Indian Council of Medical Research, New Delhi and Dept. of Health & Family Welfare, Govt. of Odisha for providing financial support for the study.

