## Supplementary file for methodology for "Natural immunity against COVID-19 significantly reduces the risk of reinfection: findings from a cohort of sero-survey participants"

A population based cross sectional serological survey carried out in August 2020 in the three largest cities of the state of Odisha in eastern India with a total population of over 2 million. The study population was randomly selected from the community members of the municipal wards from each of the city. Adults residing in the city since at least the past 3 months and who agreed to provide written informed consent for data and sample collection were included in the study. We excluded pregnant women, bed ridden patients and those with recognizable cognitive impairment. Minimum sample size per city was calculated to be 1437. This was done on the Open Epi ver3.0 software using the following formula-

*n = [Deff*Np(1-p)]/ [(d2/Z21-α/2*(N-1)+p*(1-p)]*

We assumed a sero-prevalence of 15%, which has been reported in urban regions of India during the same time period, relative precision of 20%, a design effect of 2.2 (calculated using a weak interclass correlation and a cluster size of 60), a finite population and a non-response rate of 20%. (12–14)This was rounded off to 1500 per city.

Multi-stage random sampling was used for recruiting participants. For every city, the municipal wards were treated as clusters and 25 wards were selected based on probability proportional to size. Residential street names in the ward were listed and the street from where the sampling began (as well as direction of sampling) in each cluster was selected by computerized simple random method. Households in the street were selected using systematic random sampling and one eligible individual was selected from each household using an age ordered matrix. Locked house and/or non-response were recorded and the sampling frame was shifted by one household to the immediate adjacent house in these cases. The sampling framework is provided below in figure-1.

Odisha

North- Rourkela (RKL)

25 wards 25 streets

60 HH per cluster 1 participant per HH

1500 participants

Central- Bhubaneswar (BBS)

25 wards 25 streets

60 HH per cluster 1 participant per HH

1500 participants

South- Berhampur (BAM)

25 wards 25 streets

60 HH per cluster 1 participant per HH

1500 participants

State has 3 administrative divisions- North/Central/South

Largest city from each division selected

Clusters selected based on PPS method and streets based on SRS

Households (HH) based on Systematic random sampling

Total sample size targeted was 4500 for the 3 cities

**Figure-1. Sampling framework of the Multi-stage sampling design**

Data on the socio-demographic variables, exposure history with a confirmed (and/or suspected) case, symptom profile in the last 30 days, geographical location, travel and testing history were collected in a structured tool by trained field investigators who conducted participant interviews. An Open Data Kit based electronic data capture tool was used for this purpose. Following all aseptic precautions, 3-4 ml blood samples were collected in the field by trained phlebotomists by venepuncture and transferred to vacutainers. These were transported maintaining cold chain (2-8 ^o^ C) to the serology laboratory at Indian Council of Medical Research-Regional Medical Research Centre in Bhubaneswar(ICMR-RMRC) for analysis. Additionally, secondary data on the daily number of antigen tests carried out, number of positives and deaths due to COVID-19 were obtained for the past 3 months from government sources directly.

Serum samples were subjected to detection in Roche Cobas e411 for the presence of IgG antibodies against COVID-19 using Electro-chemiluminescence immunoassay (ECLIA) based technique which is based on test principle of double-antigen sandwich assay and provides the result in 18 minutes. Elecsys® Anti-SARS-CoV-2 is an immunoassay for the in vitro qualitative detection of antibodies (including IgG) to SARS-CoV-2 in human serum and plasma. The assay uses a recombinant protein representing the nucleocapsid (N) antigen for the determination of antibodies against SARS-CoV-2. The test is intended as an aid in the determination of the immune reaction to SARS-CoV-2.

Testing procedures were followed as per the manufacturer’s instructions. Patient’s sample (20 μL) were incubated with a mix of biotinylated and ruthenylatednucleocapsid (N) antigen. Double-antigen sandwich immune complexes (DAGS) are formed in the presence of corresponding antibodies. After addition of streptavidin-coated microparticles, the DAGS complexes bind to the solid phase via interaction of biotin and streptavidin. After that the reagent mixture was transferred to the measuring cell, where the microparticles were magnetically captured onto the surface of the electrode. Unbound substances were subsequently removed. Electrochemiluminescence was then induced by applying a voltage and measured with a photomultiplier. The signal yield increased with the antibody titre. The value was expressed in Cut off Index (CoI) and a value of <1.0 was considered non-reactive and COI ≥1.0 was reactive.

The sero-prevalence of SARS-CoV-2 infection was estimated as proportion along with 95% confidence intervals and its distribution assessed across cities and demographic parameters. Gender weights were added in prevalence estimates in order to account for a higher non-response rate in females. The infection-to-case ratio and the infection fatality rate were calculated. Median time of seroconversion was assessed by a time dependent plot among those previously tested positive for SARS-CoV-2 by real time polymerase chain reaction (RT-qPCR). Temporal comparisons of the community seroprevalence estimates with the detected number of cumulative cases, active cases, recoveries and deaths was done. Heat maps for varying seroprevalence were built for each of the city’s wards. Statistical analyses were done using R (ver. 4.0.2) software packages and GIS analysis was done using QGIS (ver. 3.10).

Interviews were conducted ensuring privacy. All data was stored securely under the investigator’s responsibility, with a focus on ensuring the confidentiality of study participants. The final report and publications are based on aggregate data without any identifying information. A database with electronic tracking, password-restricted access and audit trail, with time and date stamps on data entry and edits, was used for quality control. State Health Department and city administration authorities were actively engaged to ensure smooth operationalization and also to reduce any stigma among the residents. Written informed consent for participation was obtained and a participant information sheet was provided to each household. Approval for the protocol was obtained from the ICMR RMRC Institutional Human Ethics Committee and the State Health and research ethics committee. The study methods, analyses and reporting have been informed by the WHO Unity protocols and ICMR National Serosurvey protocol in India (Kumar et al., 2020; WHO, 2020).
